# Brain Perfusion Imaging of a Large Population: Arterial Spin Labelling MRI in UK Biobank

**DOI:** 10.1101/2025.07.23.25332042

**Authors:** Thomas W. Okell, Xinyi Xu, Martin Craig, Fidel Alfaro-Almagro, David L. Thomas, Enrico De Vita, Steve Garratt, Thomas E. Nichols, Matthias Günther, Paul M. Matthews, Karla L. Miller, Stephen M. Smith, Michael A. Chappell

**Affiliations:** Oxford Centre for Integrative Neuroimaging, FMRIB, Nuffield Department of Clinical Neurosciences, University of Oxford, UK; Department of Biomedical Engineering, College of Biomedical Engineering & Instrument Science, Zhejiang University, Hangzhou, China; National Institute for Health Research (NIHR) Nottingham Biomedical Research Centre, Queens Medical Centre, Nottingham, UK; Dementia Research Centre, UCL Queen Square Institute of Neurology, UK; Dept of Translational Neuroscience and Stroke, UCL Queen Square Institute of Neurology, UK; MR Physics group, Radiology Department, Great Ormond Street Hospital, London, UK; Developmental Imaging and Biophysics Section. Developmental Neurosciences. UCL GOS Institute of Child Health, UK; UK Biobank, Stockport, UK; Big Data Institute, University of Oxford, UK; Fraunhofer Institute for Digital Medicine MEVIS, Bremen, Germany; Faculty 1, University Bremen, Bremen, Germany; mediri GmbH, Heidelberg, Germany; Department of Brain Sciences, Imperial College London; UK Dementia Research Institute Centre at Imperial College, UK; Rosalind Franklin Institute, Didcot, UK

**Keywords:** Brain perfusion, population imaging, prospective epidemiological study, arterial spin labelling, magnetic resonance imaging, cerebral blood flow

## Abstract

Blood flow to the brain is a sensitive marker of neuronal activity as well as of a number of diseases, including stroke, tumours and neurodegenerative conditions. Arterial spin labelling (ASL) is a non-invasive magnetic resonance imaging (MRI) method that can map brain perfusion, but the ability to identify relationships between blood flow and lifestyle, genetics and disease has been limited by the scale of ASL studies to date. Here, we describe the inclusion of ASL in the repeat-imaging component of the UK Biobank imaging study, a prospective epidemiological study that has acquired 100,000 first-scan datasets and aims to accumulate over 60,000 repeat-scan datasets in predominantly healthy participants, along with rich information about lifestyle factors, genetics and long-term health outcomes. The imaging protocol and analysis pipeline are outlined, along with preliminary analyses of the first 7,157 subjects (more than twice as many as the largest previous ASL study). Significant associations with a range of factors are found, including those relating to the heart and blood vessels, alcohol consumption, cognitive tasks, white matter lesions and health information, such as hearing loss and depression. ASL is shown to be more sensitive to many of these factors than other imaging modalities, complementing the existing range of structural and functional measures available in the protocol. This resource is available to researchers worldwide, which we hope will facilitate new insights into healthy brain function and pathophysiology, and potentially allow the identification of early markers of disease as long-term health outcomes accumulate.

## Introduction

Changes in brain blood flow occur in a wide range of neurovascular diseases, including vascular dementia, Alzheimer’s disease, stroke and brain tumours, and in some cases precede the appearance of clinical symptoms (Donahue et al., 2018; Gorelick et al., 2011; Luna et al., 2023). Such changes also reflect fluctuating metabolic demand in the healthy brain, indicating which regions are more or less active during a given task (Phillips et al., 2016). The ability to map brain blood flow non-invasively therefore offers the opportunity to understand healthy brain function, pathophysiology and potentially provide early markers of disease which could lead to earlier interventions and improved outcomes for patients.

Magnetic Resonance Imaging (MRI) allows brain perfusion to be measured non-invasively and without exogenous contrast agent injections using a technique known as arterial spin labelling (ASL). In this approach, inflowing blood water is magnetically labelled, allowing its accumulation in the brain tissue, and therefore regional perfusion, to be quantified (Detre et al., 1992; Williams et al., 1992). ASL has thus far mostly been used in smaller, targeted studies of specific diseases, although some larger scale applications have been undertaken, such as in the Human Connectome Project (HCP, n≈3,000 ASL datasets) (Bookheimer et al., 2019; Harms et al., 2018; Juttukonda et al., 2021; Kirk et al., 2025), the Alzheimer’s Disease Neuroimaging Initiative (ADNI, n≈1,300) (Thropp et al., 2024), the Irish National Cohort (n≈500) (Leidhin et al., 2021), and the Genetic Frontotemporal Dementia Initiative (GENFI, n≈300) (Pasternak et al., 2024). However, to disentangle complex and potentially subtle relationships between brain perfusion, lifestyle factors, genetics and pathology, much larger scale populations need to be studied and related data from detailed phenotyping, genotyping and access to health records needs to be available. In order to identify early markers of disease prior to symptom onset, such a study also needs to be prospective, with long-term access to health outcome data.

The UK Biobank imaging study (Littlejohns et al., 2020; Miller et al., 2016), part of the larger UK Biobank prospective longitudinal epidemiological study (Sudlow et al., 2015), has scanned 100,000 participants in dedicated imaging centres with identical MRI hardware and software, including brain, cardiac and body imaging protocols. Data from detailed lifestyle questionnaires, physiological measurements, genetic analysis and access to long-term healthcare records via the National Health Service are also available. Half a million middle-aged to older adults were recruited (40-69 years olds) between 2006 and 2010, to provide information on diseases and traits common in later life within a reasonable timescale. 60,000 of these subjects are being brought back for repeat imaging. Until recently, the brain imaging protocol included only structural imaging modalities (T1-weighted, T2-weighted fluid attenuated inversion recovery, susceptibility-weighted imaging), diffusion-weighted imaging and task and resting state functional MRI, none of which provide direct insights into brain physiology.

Here we describe the recent addition of ASL into the UK Biobank imaging protocol. This will be performed on all subjects during their repeat visits, aiming for the acquisition of over 60,000 second-scan datasets by the end of 2028. The ASL protocol and processing pipeline are described along with some preliminary analyses of over 7,000 subjects, to demonstrate the potential of this dataset. These data will provide a new data resource in UK Biobank with which researchers can explore detailed relationships between brain perfusion and lifestyle factors, genetics and disease processes, including multi-organ comorbidities.

## Results

### Overview of the ASL protocol and analysis pipeline

The duration of the UK Biobank brain imaging protocol is limited to ∼35 minutes in order to maintain a sufficiently high throughput of participants at the four dedicated imaging centres to allow completion of scanning of these large cohorts. The addition of ASL became possible in the repeat scan protocol by shortening the duration of the task functional MRI (fMRI) protocol from 4 to 2 minutes, opening up a 2-minute window. While this is shorter than is allowed for a typical ASL scan (∼5 minutes), we found that it was sufficient to generate robust data after careful protocol optimisation.

After piloting, results suggested that the optimal protocol for this study would be multiple postlabelling delay (PLD) pseudocontinuous ASL (PCASL) (Dai et al., 2008) with background suppression (Ye et al., 2000) and a segmented 3D gradient and spin echo (3D-GRASE) readout (Günther et al., 2005). This protocol provided an acceptable balance between imaging speed, signal-to-noise ratio, motion sensitivity and resolution (see the Methods section for further discussion). The use of a multi-PLD approach allows the simultaneous estimation of quantitative tissue perfusion and arterial transit time, maintaining robustness to delays in blood arrival across brain regions and subjects. Arterial transit time is also an interesting physiological parameter in its own right (MacIntosh et al., 2010). This protocol was then incorporated into the UK Biobank brain imaging study. Here we will describe results from ASL on the first 7,157 subjects studied.

A robust, automated post-processing pipeline was also developed for this project. An overview of the acquired ASL data and image analysis pipeline is given in **Figure 1**. Briefly, the acquired ASL data were motion and distortion corrected, pairs of images subtracted to isolate the perfusion-weighted signal, kinetic model fitting was performed and the signal calibrated to produce maps of absolute perfusion (Cerebral Blood Flow, CBF) and arrival delay (Arterial Transit Time, ATT) for each subject. These maps were also non-linearly registered to a standard template to allow comparison across subjects. Further details of the protocol and analysis pipeline are given in the Methods section.

**Figure 1:**
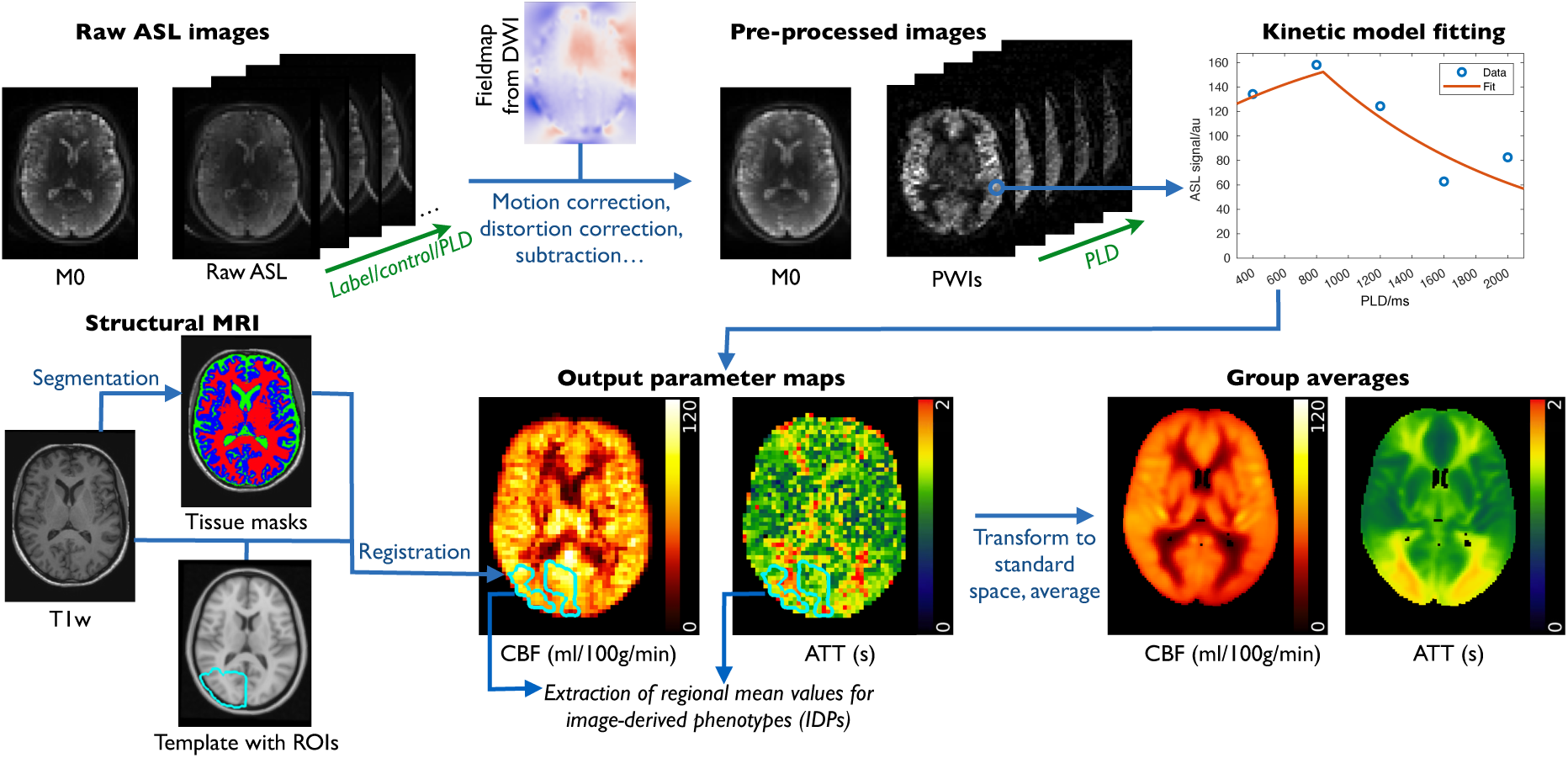
Overview of the UK biobank ASL data and processing pipeline. The raw calibration (M0) and label/control images at each postlabelling delay (PLD) are pre-processed to correct for motion, B_0_-induced distortion and gradient non-linearities and the ASL data are control-label subtracted to give corrected M0 and perfusion-weighted images (PWIs) at each PLD. Kinetic model fitting in each voxel and calibration with the M0 image gives an estimate of absolute cerebral blood flow (CBF) and arterial transit time (ATT) in each voxel. The ASL data are registered to the structural T1-weighted images processed by the original UK biobank pipeline, which include the necessary warp fields from standard (MNI) space, allowing tissue-type masks and regions of interest to be overlaid on the ASL maps and regional mean values to be extracted as Image-Derived Phenotypes (IDPs). The CBF and ATT maps from each subject are also registered to standard space to allow voxelwise cross-subject analyses to be performed and group average templates to be derived.

### Image-Derived Phenotype (IDP) generation

Summary measures from the CBF and ATT maps of each subject were calculated as mean values within various regions of interest (ROIs), including major brain lobes, subcortical structures and vascular territories. Many of these were confined to grey matter, for which the ASL signal is more reliable, although ROIs for some larger white matter regions were also included. This resulted in 50 ASL-based image-derived phenotypes (IDPs) per subject.

Deconfounding of the IDPs and voxelwise maps was performed (Alfaro-Almagro et al., 2021) to remove the effects of factors such as age, sex, imaging site and head size, but also haematocrit (known to influence the ASL signal via blood relaxation rates (Lu et al., 2004)) and cortical thickness (to mitigate partial volume effects).

### Data quality and simple associations

The expected strong trends of decreasing CBF and increasing ATT with age (**Figure 2**A) and higher CBF in females than males (**Figure 2**D) were robustly demonstrated in this cohort (prior to deconfounding), despite the short scan time available for each subject. The data quality was also demonstrated by the strong correlations with age visible at the voxel level across most of the brain (**Figure 2**B) and the ability to see more subtle variations, for example with reducing CBF and increasing ATT throughout the day (**Figure 2**C), in line with previous observations (Hodkinson et al., 2014).

**Figure 2:**
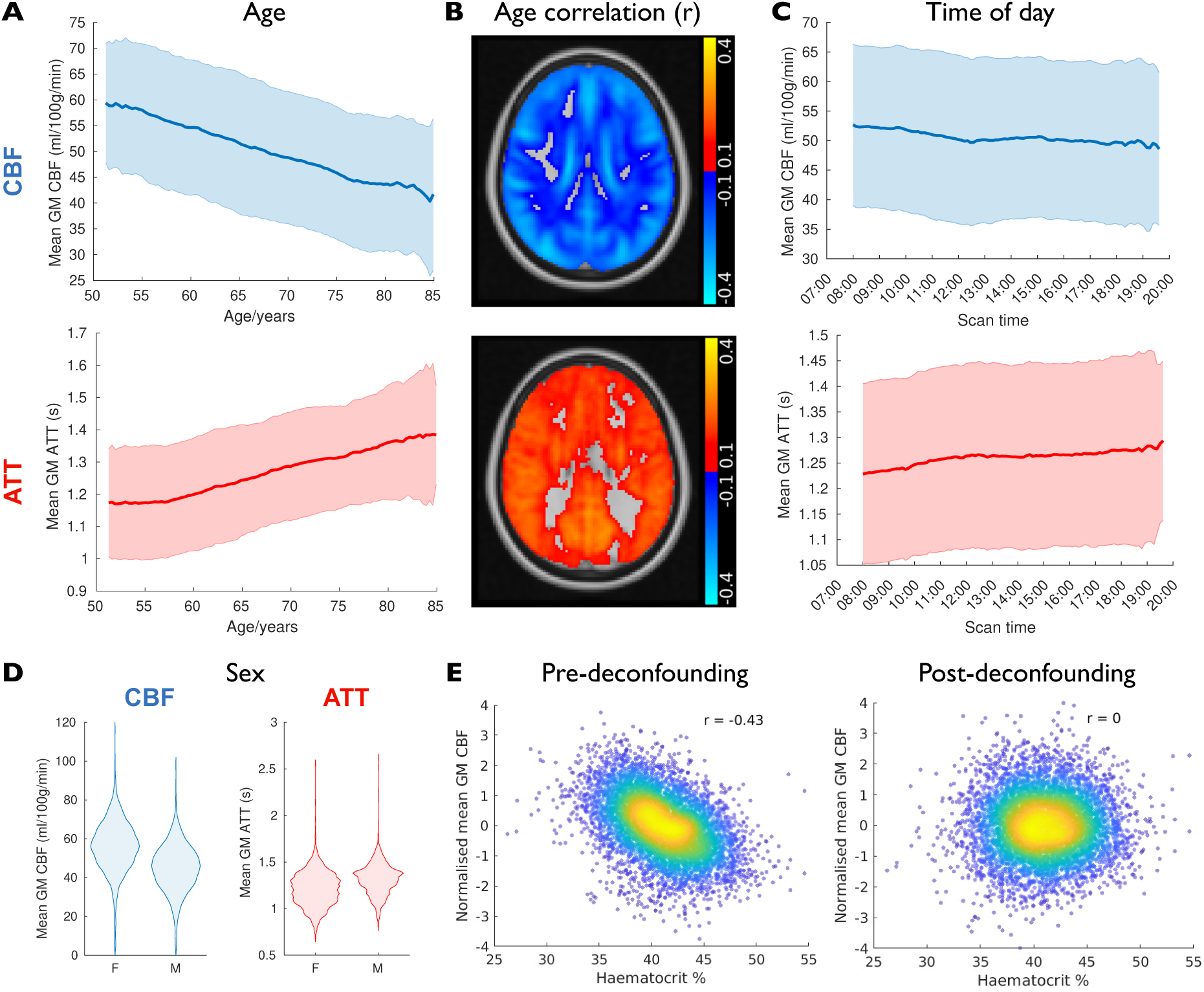
Simple associations and deconfounding. Despite the short scan time available for each subject, the mean grey matter (GM) CBF shows the expected robust decrease with age, along with a concomitant increase in ATT (shown as a sliding window mean and standard deviation in **A**, window width 5 years). This robust correlation is also present at the voxelwise level (**B**), with most of the brain showing strong correlations of CBF and ATT with age (shown here as maps of the Pearson correlation coefficient, *r*, in standard space). Small variations across the day (**C**, sliding window width 2.6 hours) and with sex (**D**) are also evident. The effect of haematocrit on the ASL IDPs is significant (**E**), but is removed by the deconfounding process.

The expected strong sensitivity to haematocrit was also observed before being removed by deconfounding (**Figure 2**E), highlighting the importance of regressing out confounds before exploring associations with other factors. This is especially important in such a study with a large number of subjects where even subtle (valid or artefactual) associations can be detected. Deconfounding should limit potentially misleading associations between, for example, brain perfusion and a dietary factor, where this relationship is entirely mediated by haematocrit.

### Arterial transit time distributions and postlabelling delay suggestions

The large quantity of multi-PLD ASL data collected here allows exploration of the ATT distribution across the population (**Figure 3**). This has implications for single PLD ASL acquisitions, where the PLD needs to exceed the largest ATT present in every subject to avoid CBF underestimation, but excessively long PLDs lead to weaker perfusion signals. Understanding the transit time distribution is also useful for designing optimized multi-PLD protocols (Woods et al., 2024, 2019). These distributions are approximately consistent with the previous single PLD recommendation of 2 s in elderly subjects (Alsop et al., 2015), which exceeds the ATT in more than 98% of grey matter voxels in this cohort, although a PLD of up to 2.5 s would be needed to accurately estimate CBF in 99% of grey matter voxels in 99% of subjects, particularly in older people. However, this comes with the caveat that the multi-PLD protocol used here is not sensitive to extremely delayed blood arrival (ATT > 3.8 s).

**Figure 3:**
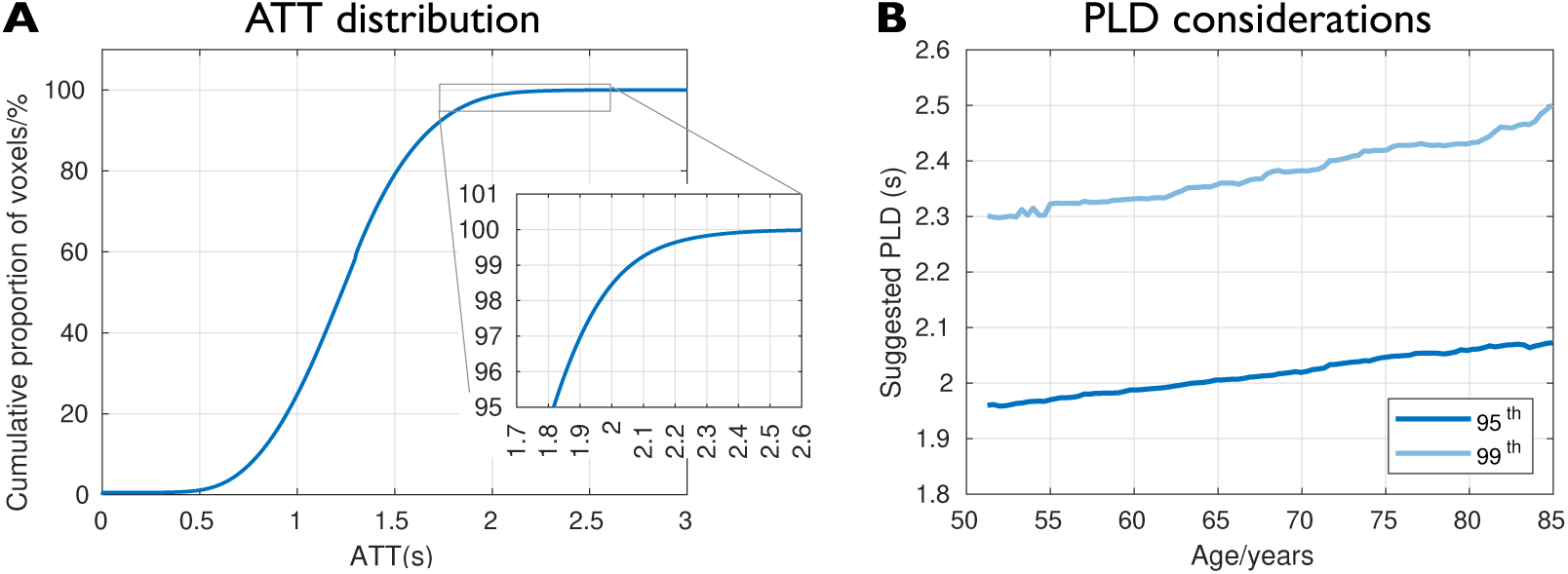
Arterial transit times and postlabelling delay considerations. **A**) The cumulative distribution of arterial transit times (ATTs) across all grey matter voxels in all subjects shows that more than 98% of voxels have ATTs of 2s or less in this cohort. **B**) These distributions can also be used to calculate the 95^th^ or 99^th^ percentile ATT values in grey matter for each subject. The 95^th^ or 99^th^ percentiles of these values in a sliding window across ages (window width 5 years) is plotted here, suggesting the postlabelling delay (PLD) necessary to accurately estimate CBF with single-PLD ASL in this proportion of voxels and subjects.

### Associations between ASL and other measures

After IDP generation and deconfounding, we calculated univariate associations between ASL IDPs and the thousands of lifestyle, health and physiological measures present in the UK biobank. A Manhattan plot showing the significance of these associations, grouped by category, is shown in **Figure 4**A. Even after deconfounding and correcting for multiple comparisons, over 150 non-imaging measures were found to be significantly associated with at least one ASL IDP (a total of 1,439 associations passed the false discovery rate threshold, with the most significant for each non-imaging measure listed in Supplementary Table 1). Interestingly, 48% of these non-imaging factors were more strongly associated with ATT than CBF, showing that ATT provides complementary information to flow and highlighting an additional advantage of using a multi-PLD ASL approach.

**Figure 4:**
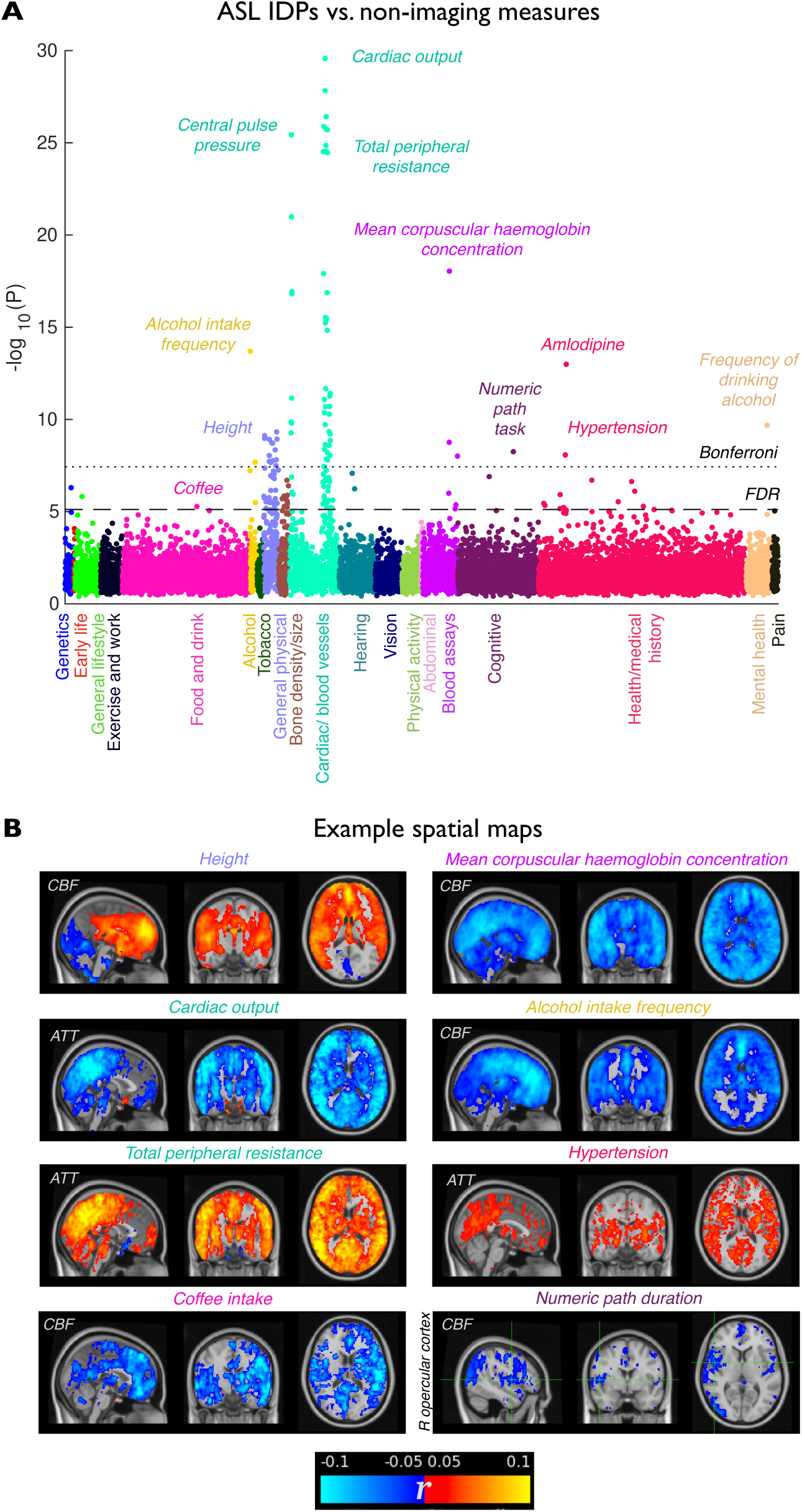
Correlations between ASL and other (non-imaging) measures. **A**) Manhattan plot showing the significance of univariate associations between ASL IDPs and all the non-imaging variables grouped and colour-coded by category along the x-axis. Each dot represents the most significant association with an ASL IDP, plotted as −log_10_(P) on the y-axis, with larger values being more significant. Bonferroni and false discovery rate (FDR) multiple comparison correction thresholds are also shown. Some interesting associations exceeding the multiple comparisons thresholds are highlighted. **B**) Spatial correlation maps corresponding to selected significant associations with non-imaging variables (shown as Pearson’s correlation coefficient maps, *r,* in standard space with positive correlations in red-yellow and negative in blue-light blue, overlaid on the MNI template using radiological convention L=R). Some of these factors have a near-global effect on CBF or ATT, while others are more spatially specific, such as the lower CBF in frontal regions associated with alcohol intake, ATT increases in deep grey matter with hypertension and CBF reductions particularly in right opercular cortex associated with poorer performance in a cognitive task (numeric path duration).

Many of these significant associations are with large scale physical, cardiac and blood vessel factors, but there are a number of other categories with significant relationships, including lifestyle, food and drink, cognitive performance and health data. These factors can be regressed back into the voxelwise perfusion data to generate spatial maps of these associations that can aid interpretation (Figure 4B) and help identify cases where the relationships may be artefactual (some examples are given in Supplementary Figure 1).

These spatial maps revealed that some non-imaging measures are associated with largely global perfusion differences. Physiological hypotheses can be generated for some of these relationships: for example, higher cardiac output is associated with shorter ATTs (likely driven by the more rapid blood movement from the heart to the brain), higher peripheral resistance is associated with longer ATTs (increased vascular resistance would be expected to lead to slower blood arrival) and higher mean corpuscular haemoglobin is associated with reduced CBF (since each red blood cell can deliver more oxygen to the tissue (Van Der Veen et al., 2015)). Subject height may influence the way blood flow is regulated in individuals of different sizes, particularly whilst lying supine in a scanner, with larger increases in flow in the anterior vs. posterior circulation previously observed when moving from a seated to a supine posture (Possnig et al., 2025). Coffee intake (self-reported cups per day) was also associated with lower CBF in most brain regions, most likely due to the acute pharmacodynamic effects of caffeine on cardiac and vascular functions (Addicott et al., 2009).

Others associations were more spatially specific: for example, alcohol intake is linked to reduced CBF throughout the brain but it is particularly highly correlated in frontal regions known to be affected in chronic alcoholism (Kuruoglu et al., 1996); hypertension is associated with longer ATTs, particularly in deep grey matter regions, which are supplied by the deep, small penetrating arteries prone to hypertension associated arteriosclerosis (Hainsworth et al., 2024); and the time taken to complete the numeric path cognitive test corresponds to reduced CBF in specific regions including the right opercular cortex, a region known to relate to cognitive control of response selection (Higo et al., 2011).

### ASL sensitivity

Although we found significant associations between ASL IDPs and many non-imaging measures, it is possible that other structural or functional brain imaging modalities could provide comparable information. To explore whether ASL is more sensitive than other imaging modalities, the significance of associations between groups of IDPs from different modalities were compared to ASL for each non-imaging measure (see the Methods section for details). Of the 153 non-imaging measures identified above, ASL had significantly stronger associations compared to other modalities in 69 (45%) of them (shown by the points exceeding the false discovery rate threshold in **Figure 5**A, which are also listed in Supplementary Table 2), demonstrating that ASL is providing additional information over and above other modalities.

**Figure 5:**
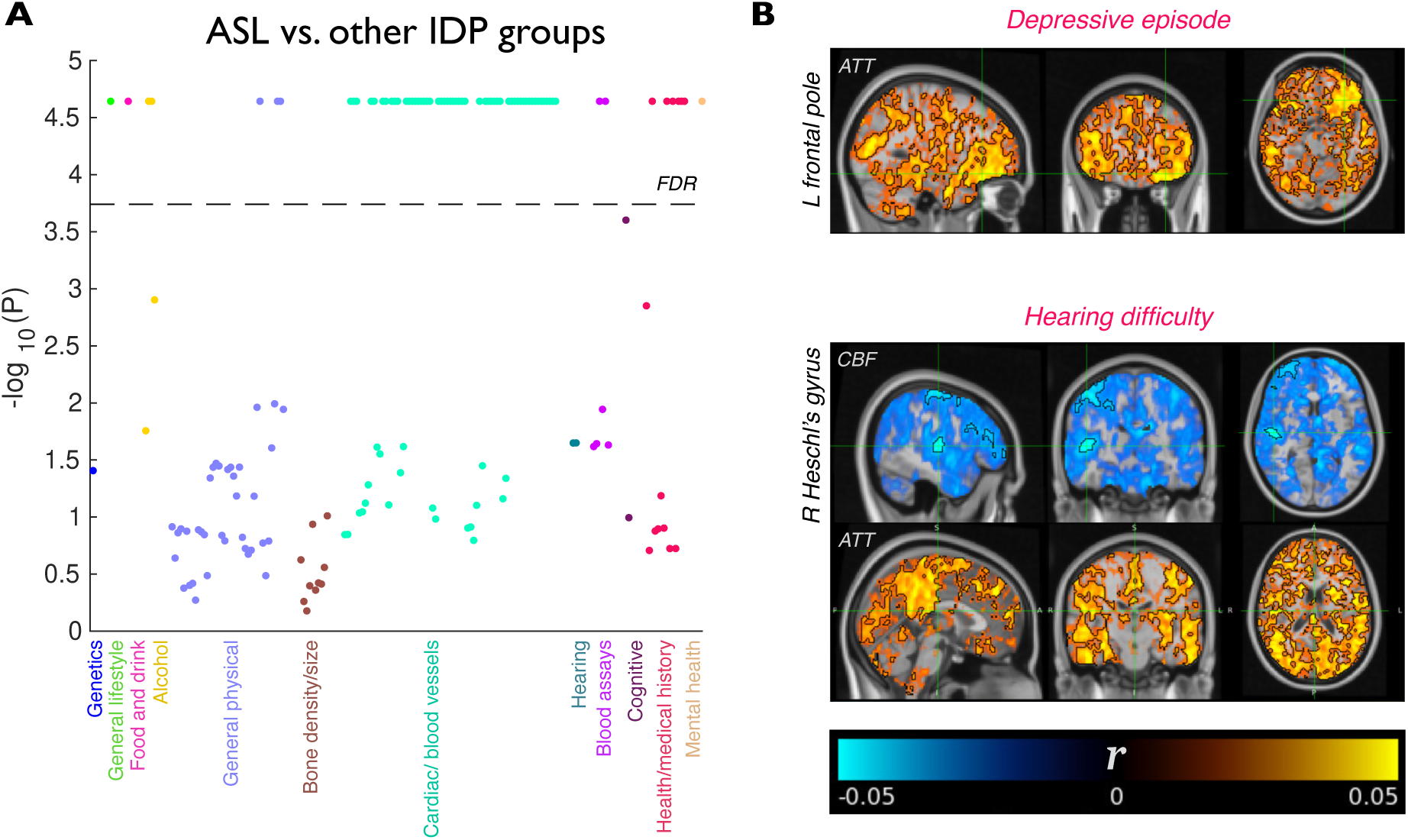
The sensitivity of ASL to other (non-imaging) measures. **A**) Manhattan plot showing the strength of associations between non-imaging measures and the ASL IDP group relative to similarly sized groups of IDPs from other modalities obtained from permutation testing (see Methods for details). −log_10_(P) values exceeding the FDR threshold indicate that ASL is significantly more strongly associated with this non-imaging measure than other modalities. Note that the maximum −log_10_(P) value achievable is limited by the number of IDP groups and permutations used. All non-imaging measures were included in statistical testing, but only those which were significantly correlated with at least one ASL IDP are shown here for clarity. **B**) Two examples of health-related measures where ASL was more sensitive than other modalities. Subjects who have suffered from a depressive episode had significantly longer ATTs, particularly in the left frontal pole, as well as sub-threshold CBF reductions in a number of other brain regions (see Supplementary Figure 2). Hearing difficulty was significantly associated with reduced CBF in several auditory processing regions including right Heschl’s gyrus, precentral cortex and frontal regions, as well as widespread ATT increases. To show these subtle associations more clearly, the statistically significant regions are highlighted with a black outline and overlaid on the spatial correlation maps, with opacity modulated by the correlation coefficient, as recommended by (Taylor et al., 2023).

We found that ASL is particularly sensitive to some measures of health: for example, subjects who have experienced a depressive episode have longer ATTs, particularly in the left frontal pole, a region previously reported to be affected in depression (Bludau et al., 2016; Veer, 2010). The ATT relationship is significant (**Figure 5**B) and there are sub-threshold CBF reductions in other brain regions previously found to be affected in depression (Supplementary Figure 2).

ASL was also particularly sensitive to hearing loss: CBF reductions were found in Heschl’s gyrus, a primary region for auditory processing, as well as in functionally downstream regions in precentral cortex and the frontal lobe (Hakonen et al., 2017). The more significant hearing difficulty association in the right hemisphere found here has been reported previously (Ponticorvo et al., 2019). This either may reflect differences in the role of the right Heschl’s gyrus (e.g., arising from its specialisation for frequency-specific processing (Warrier et al., 2009)) or because this region is more anatomically consistent across subjects in the right hemisphere (Ren et al., 2021), making associations easier to detect. Widespread ATT increases were also found to be associated with hearing loss.

### Correlations with other imaging modalities

The simultaneous acquisition of ASL data with a number of other brain imaging modalities in this large cohort provides an opportunity to explore the relationships between them. The ASL IDPs were strongly associated with many IDPs from other brain imaging modalities (1,772 significant correlations with at least one ASL IDP, listed in Supplementary Table 3), particularly with regional/tissue volumes, tissue intensities, T2* measures, Quantitative Susceptibility Mapping (QSM), white matter Diffusion Tensor Imaging (DTI) metrics and resting fMRI node amplitudes (**Figure 6**).

**Figure 6:**
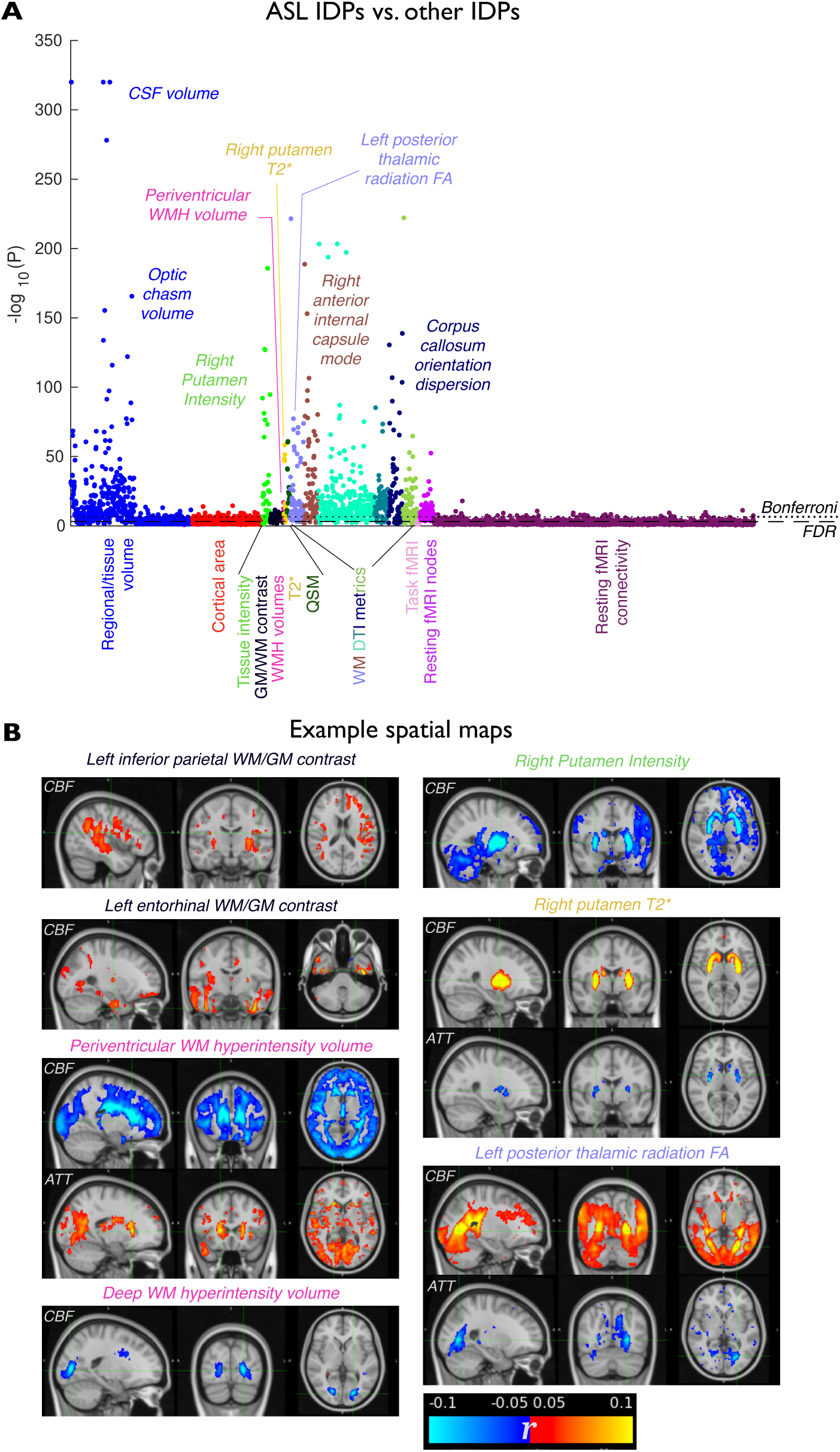
Correlations between ASL and other brain imaging measures. **A**) Manhattan plot of the univariate associations between ASL IDPs and all other brain imaging IDPs, grouped and coloured by modality along the x-axis. Many associations exceed the Bonferroni and FDR thresholds for significance, with a few non-ASL IDPs of interest highlighted here. **B**) Voxelwise spatial maps of correlation coefficients between CBF/ATT and some selected non-ASL IDPs. These include increased CBF in regions with higher white matter/grey matter contrast, reduced CBF and increased ATT where more white matter hyperintensities are found, associations with deep grey matter T1-weighted intensity and T2*, and even increased CBF and reduced ATT in white matter regions associated with higher fractional anisotropy (FA, derived from diffusion tensor imaging).

The spatial maps of these correlations reveal some interesting patterns. For example, white/grey matter contrast in the inferior parietal lobe is associated with higher CBF in the same region, but also in deep grey matter regions such as the putamen. Similarly, higher contrast in entorhinal cortex correlates with higher CBF in this region as well as ventral temporal areas. This higher contrast may reflect “healthier” brain tissue (Jefferson et al., 2015; Salat et al., 2009) which has a higher metabolic demand and this has a downstream effect on other regions it is strongly connected to.

A greater volume of white matter hyperintensities (lesions), which arise with aging and with neurodegenerative diseases (Dadar et al., 2022), correlates with reduced CBF in the same regions. This demonstrates the potential for sensitivity to white matter perfusion changes with our protocol in this large cohort, despite the challenges of using ASL in these regions (van Gelderen et al., 2008). Although there may be coincident brain atrophy leading to partial volume contributions to the observed periventricular CBF changes, the region of CBF reductions is well beyond the ventricular boundaries and there is a concordant increase in ATT, which is less sensitive to the confounds of partial volume. Similar changes are also seen in the deep white matter, where partial volume effects are less likely to occur. CBF and ATT correlation maps also coincide with DTI metrics in white matter regions such as the posterior thalamic radiation, with higher CBF being associated with higher fractional anisotropy, another measure thought to be sensitive to tissue integrity (Raghavan et al., 2020).

Strong correlations with deep grey matter signal intensities (on T1w imaging) and T2* were also found. Although there is the potential for direct signal modulation caused by changes in tissue relaxation properties affecting CBF estimates, the use of voxelwise calibration in our analysis pipeline should remove these effects. These observations perhaps therefore arise from the relationship between iron accumulation in deep grey matter regions and tissue health (Madden and Merenstein, 2023), driving, for example, the correlation between longer T2* (lower iron accumulation) and higher CBF (more metabolically active tissue).

Strong associations between ASL and resting-state fMRI IDPs were also observed. Here we provide only a preliminary exploration of these complex relationships and have limited our analyses to the stronger associations between ASL and rfMRI node (region) amplitudes rather than connectivity. The spatial maps of associations between CBF and rfMRI node amplitudes (**Figure 7**) show stronger correlations within regions associated with a given node, and/or more negative correlations outside of these regions. Indeed, the average correlation coefficient inside each node mask is significantly larger than that outside (P = 0.007). We speculate that this may be driven by a complex interplay between, e.g.,: a) a higher metabolic demand and therefore higher CBF in the associated rfMRI node regions, possibly at a cost to blood flow in other brain regions; b) the potential dampening of BOLD signal fluctuations when baseline CBF is higher (Stefanovic et al., 2006); and c) other confounding factors, particularly in subjects with compromised neurovascular coupling (Amemiya et al., 2012). However, further work is needed to unravel this complex relationship.

**Figure 7:**
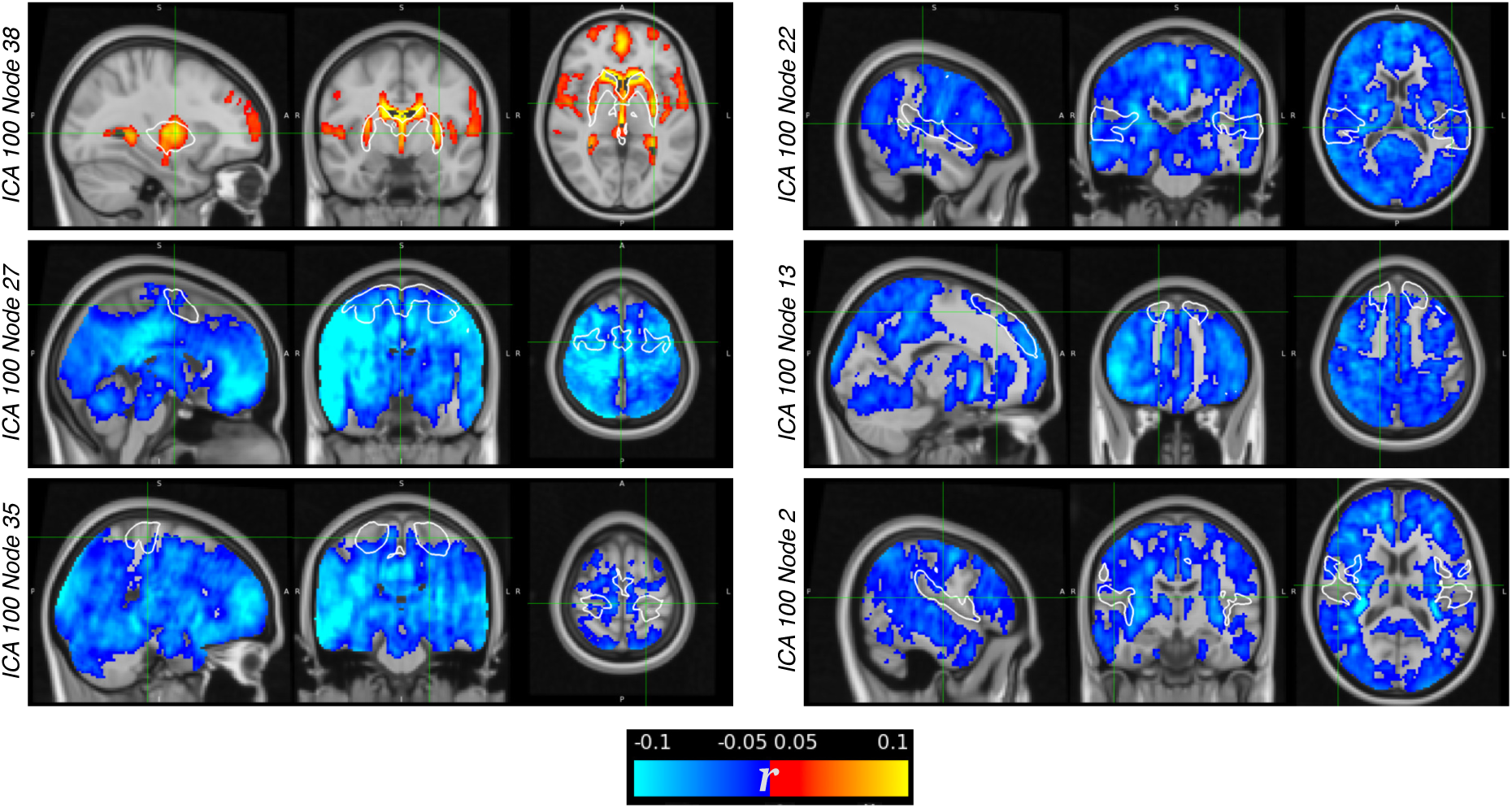
Associations between ASL and resting fMRI. Correlation coefficient maps of CBF against selected node amplitudes are shown in order of decreasing significance (top to bottom and left to right). The regions associated with each node are highlighted with a white outline (z > 5). It appears that higher node amplitudes generally correspond to higher CBF within the node region, or reduced CBF outside that region.

Finally, spatial correlation maps may also help to identify significant relationships that are likely driven by artefacts. For example, many of the volumetric IDPs strongly correlate with CBF, but these associations are likely driven by partial volume effects or the influence of arterial signals on structural imaging IDPs (some examples are given in Supplementary Figure 1).

## Discussion

### Perfusion imaging in UK Biobank

The introduction of ASL into the UK Biobank brain imaging study represents the first direct physiological measure available in this protocol, supplementing the existing structural and functional information which has already provided exciting new insights in a range of applications, e.g. (Cox et al., 2019; Elliott et al., 2018; Miller et al., 2016; Wang et al., 2022). As far as we are aware, the 7,157 datasets presented in this preliminary study is more than twice the size of the largest existing ASL study (∼3,000 subjects in the Human Connectome Project (Bookheimer et al., 2019; Juttukonda et al., 2021; Kirk et al., 2025)), and will be an order of magnitude larger (60,000 subjects) once data collection is complete. This opens up a myriad of possible investigations into the relationships between brain perfusion measures and lifestyle factors, genetics and disease processes that have not been possible previously, aided by the rich phenotypic, genetic and health outcome data available in UK Biobank.

Despite the short scan time available, the robust correlations between ASL measures and known factors, such as sex, time-of-day and age, even at the voxel level, suggest that the data quality is sufficient to retain sensitivity to a wide range of physiological differences. The large scale of this study and the use of a multi-PLD protocol allows the ATT distribution in this cohort to be robustly defined, providing guidance for others designing ASL protocols in other studies. Whilst longer ATTs were seen in white matter regions (e.g. see the group average in Figure 1), we caution against a strong interpretation of the white matter ATT distributions derived here since the range of PLDs used in this study, along with the inherently low signal-to-noise ratio of the data, may have been insufficient to reliably estimate ATT in some white matter regions (van Gelderen et al., 2008). Through-slice blurring effects could also contaminate the weak white matter signal with stronger nearby grey matter signals, biasing the derived distributions.

The sensitivity of the derived ASL metrics is also demonstrated by the wide range of strong associations with imaging and non-imaging measures found in this initial study. As well as the expected associations with cardiac and physical factors, more subtle associations could also be identified, such as the reduction in frontal lobe CBF associated with alcohol intake, the change in perfusion in brain regions responsible for auditory processing in those with hearing loss, perfusion changes in those who have experienced a depressive episode and even higher white matter CBF in white matter tracts with higher fractional anisotropy (often thought of as a proxy for “structural integrity”). These are only a small fraction of the associations found in this preliminary study. The broad phenotypic information available in UK Biobank will allow such relationships to be studied in greater detail in follow-on studies, particularly as the number of datasets continues to grow, providing even greater statistical power.

### Challenges

The two minutes of imaging time available for ASL is very limited, meaning that the individual data from each subject are relatively noisy. This may have contributed to the relatively small correlation coefficients found in univariate analyses with other imaging and non-imaging variables, although other UK Biobank imaging studies have reported comparable levels of association (Miller et al., 2016; Wang et al., 2022), suggesting this is a common feature in similarly sized imaging studies. Nevertheless, the statistical power afforded by the large number of subjects allows such correlations to reach significance, underlining the need for careful deconfounding and assessments for meaningfulness, such as through the use of spatial maps to identify potentially artefactual correlations. Many of the associations found here were interpretable and consistent with previous literature, but as has been noted previously (Miller et al., 2016; Smith and Nichols, 2018), care must be taken to avoid interpreting the associations as causal, since similar variation in two measures could be driven indirectly by a separate factor. Combining ASL with other imaging and non-imaging measures in multi-variate analyses could help with interpretability and explain a greater proportion of the variance in the data in future studies (Miller et al., 2016).

Another compromise made in the protocol design was that between spatial resolution and sensitivity. The relatively coarse voxel size (3.4×3.4×4.5 mm), compounded by the through-slice blurring effect from the 3D-GRASE readout (Günther et al., 2005), led to significant partial volume effects. This was mitigated here by the inclusion of cortical thickness in the deconfounding process, although a region-specific correction approach that is aware of the through-slice blurring (Boscolo Galazzo et al., 2014) is desirable in future iterations of the ASL analysis pipeline. The largest previous ASL study, the Human Connectome Project (HCP), made different protocol choices, using a simultaneous multi-slice readout that does not suffer from through-slice blurring, and a relatively high spatial resolution, but without background suppression (Bookheimer et al., 2019), favouring spatial fidelity over SNR efficiency. Combining the relative advantages of both datasets could be an exciting avenue of future research.

The large size of the UK Biobank dataset precludes fully manual quality control. In this study, subjects were excluded if the subject’s T1-weighted structural image was deemed to be of insufficient quality (Alfaro-Almagro et al., 2018), which prevents accurate tissue-type segmentation and registration, and motion-related parameters derived from resting-state fMRI were included as potential confounding factors. However, as per other most other imaging modalities, no quality control measures were directly derived from the ASL data. Automated quality control for ASL may therefore be beneficial in the future (Dolui et al., 2024).

This dataset could provide useful normative brain perfusion distributions for comparison with other studies. However, quantitative harmonization of brain imaging data is challenging (Pomponio et al., 2020), and this is likely to be even more difficult for ASL data, due to the wide range of ASL labelling strategies, timings, background suppression methods, readout approaches (which influence the point spread function) and processing pipelines that are used (Hernandez-Garcia et al., 2022), as well as the usual variations found across scanner manufacturers, hardware and software versions. Much work in this field is ongoing (Warrington et al., 2023), but additional studies focused on ASL would be beneficial.

### Future prospects

As the number of ASL datasets accumulates, this resource will provide increased statistical power to explore subtle perfusion differences across subjects and their relationships with lifestyle and health factors. It will also become feasible to explore genetic associations with brain perfusion, which could have implications for a range of cerebrovascular diseases by, for example, identifying new targets that could modulate brain blood flow.

As time progresses, additional health outcomes will begin to accumulate, such as Alzheimer’s disease, strokes and vascular dementia, allowing researchers to look back to UK biobank perfusion data acquired prior to diagnosis and retrospectively identify markers of clinical disease risk that could guide prospective studies for earlier diagnosis and intervention in the future. ASL is also a sensitive marker of brain age, as shown here and in other studies (Dijsselhof et al., 2023). Combining ASL with other features from structural, diffusion and functional imaging could potentially improve the sensitivity of brain age deltas (the difference between a subject’s real and predicted age) to various disease processes (Smith et al., 2019). Here we presented some preliminary findings on links between perfusion and resting-state fMRI, but clearly this is a complex relationship, both in terms of the mechanisms driving the fMRI contrast (Blockley et al., 2013) and the underlying associations between neural activity and metabolism, both in health and disease. This large dataset will help researchers to explore these relationships in greater detail and could, for example, lead to methods to remove aspects of the resting fMRI signals that are driven by physiological processes rather than neural activity.

The UK Biobank perfusion imaging resource complements the existing imaging modalities and provides a wide range of opportunities for researchers to explore novel links between brain blood flow and a range of other lifestyle, genetic and health outcome data available in this large study.

## Methods

### Ethics

Ethical approval for the UK Biobank study to acquire and disseminate data and samples from the participants was obtained through the North West Multi–centre Research Ethics Committee, and these ethical regulations cover the work in this study (http://www.ukbiobank.ac.uk/ethics/). All participants provided written informed consent.

### Subject demographics

The UK Biobank study recruited 500,000 participants aged between 40 and 69 years at baseline (Sudlow et al., 2015). 100,000 of these have been scanned for the imaging study (Miller et al., 2016) and 60,000 of these will return for repeat imaging, where ASL is included in the imaging protocol. In this work, the 7,157 participants from the October 2023 data release who had ASL data and T1-weighted imaging that passed quality control (Alfaro-Almagro et al., 2018) were included. Participants’ mean age at the imaging visit was 66.8 ± 7.7 years (range 51-84) and 53.8% were female.

### Imaging Protocol

The original UK Biobank brain imaging protocol including structural, diffusion and functional MRI has been detailed previously (Miller et al., 2016). Scanning was performed on 3T Siemens Skyra systems (software version VD13D) with a 32-channel receive head coil, at four dedicated imaging sites. In the latest version of the protocol, task fMRI was cut down from 4 to 2 minutes, allowing a 2-minute ASL protocol to be added without extending the total scan time. Since ASL is an intrinsically low signal-to-noise ratio (SNR) technique, the protocol choices had to carefully balance SNR, spatial resolution, blurring, motion robustness and sensitivity to a range of arterial transit times within this short acquisition window.

After careful piloting, we arrived at a protocol with the following key features: 1) a 1.8 s balanced pseudocontinuous ASL (PCASL) preparation (Dai et al., 2008) to label the inflowing blood, as recommended previously (Alsop et al., 2015), with the labelling plane parallel and 90 mm inferior to the centre of the imaging volume; 2) a background suppression scheme to minimize physiological noise and sensitivity to subject motion (Ye et al., 2000), consisting of a pre-saturation module and two global inversion pulses timed to null tissues with a T1 of 700 ms and 1400 ms (Günther et al., 2005), approximately corresponding to the values for grey and white matter, at a time 100 ms before the readout; 3) five post labelling delays (PLDs), ranging from 400 ms to 2000 ms in steps of 400 ms, to better quantify cerebral blood flow (CBF) in the presence of varied arterial transit time (ATT: the time taken for blood to travel from the labelling plane to the brain tissue) across brain regions and individuals, and further to allow the ATT also to be estimated as an interesting parameter in its own right, with the repetition time minimized for each PLD separately; 4) an SNR-efficient 2-shot 3D gradient and spin echo (3D-GRASE) readout (Feinberg and Oshio, 1991; Günther et al., 2005) with left-right phase encoding, 29 ms echo time, voxel size 3.4×3.4×4.5 mm, 32 slices, 120° refocusing pulse flip angle and 6/8^th^ slice partial Fourier (resulting in a turbo factor of 12) to minimize the readout duration, giving a good compromise between imaging speed, motion sensitivity, through-slice blurring and background suppression efficiency; and 5) a calibration (M0) image without PCASL labelling or background suppression and a long repetition time of 5390ms, to allow CBF to be quantified in absolute units (ml blood/100g tissue/minute).

This resulted in an acquisition time of exactly 2 minutes. Unlike the fMRI and DTI protocols, which used strong slice angulation to minimize the number of slices required to cover the brain, the ASL protocol used transverse/axial slice orientation to prevent the labelling plane (automatically positioned parallel to the imaging region) from intersecting the brain feeding arteries too far from isocentre where B_0_ homogeneity is poorer, leading to reduced PCASL labelling efficiency in preliminary testing. To accommodate this orientation, a larger 144 mm field of view was used in the slice direction, which should encompass the whole brain in ∼99.5% of subjects or more, given the reduction in brain size with age (Mennes et al., 2014). A copy of the full protocol is available online: https://www.fmrib.ox.ac.uk/ukbiobank/protocol/index.html.

### Image processing pipeline

A robust, automated post-processing pipeline was also developed based on the BASIL toolbox (Chappell et al., 2023), part of the FMRIB Software Library (FSL) (Jenkinson et al., 2012). Pre-processing included correction for gradient non-linearities (Alfaro-Almagro et al., 2018), subject motion (Jenkinson, 2002) and B_0_-induced distortions (using a fieldmap derived from the blip-up/down diffusion data (Andersson et al., 2003)). Pairs of control-label images were then subtracted to isolate the perfusion-weighted signal at each PLD.

Fitting to the general ASL kinetic model (Buxton et al., 1998) was performed using a variational Bayesian framework (Chappell et al., 2009), incorporating a macrovascular component (Chappell et al., 2010). The signal was then voxelwise calibrated to produce maps of absolute CBF and ATT for each subject, along with voxelwise estimates of parameter precision. Partial volume correction (Chappell et al., 2011) was not applied in this pipeline due to the additional complication of through-slice blurring from the 3D-GRASE readout. We aim to develop and incorporate a blurring-aware partial volume correction in future versions of the analysis pipeline.

Alignment between the pre-processed ASL data and the subject’s T1-weighted anatomical image was achieved using linear registration (Jenkinson, 2002), and brain-boundary registration (Greve and Fischl, 2009) using the correspondence between the control-label subtraction image averaged across PLDs (which has better grey-white matter contrast than the original ASL data) and the T1-weighted structural. Non-linear registration (Andersson et al., 2007) of each subject’s T1-weighted structural image to the MNI152 template was previously performed (Alfaro-Almagro et al., 2018), allowing all CBF and ATT maps to be transformed into standard space for between-subject analyses.

### Image-Derived Phenotype generation

Regions of interest (ROIs) for IDP generation were defined either in standard space or each subject’s structural space and warped back to the native space of the ASL data before precision-weighted mean values (i.e. upweighting those voxels where the fitting procedure had more confidence in the estimated parameters) for CBF and ATT were extracted as IDPs. Grey matter ROIs included the whole brain, the cortex, separate right and left hemisphere major brain lobes and the cerebellum (defined in standard space from the MNI atlas), and the major cerebral vascular territories of the right and left internal carotid arteries and vertebrobasilar arteries (excluding the cerebellum). Canonical vascular territory ROIs were generated based on an MNI template (Mutsaerts, 2015) derived from previous work (Tatu et al., 1998), with the anterior and middle cerebral artery masks merged to form internal carotid artery masks. To improve the separation of signals from different vascular territories and account for some of the variability in these regions across subjects, these masks were eroded first by a 3×3×3 kernel and again by a 5×5×5 kernel (at 2 mm isotropic resolution), but only removing voxels if another vascular territory was present within the kernel. All grey matter ROIs were further refined using a subject-specific grey matter mask (defined as having more than 70% grey matter within a given voxel). Caudate, putamen and thalamus masks were also derived from the segmentation of each subject’s structural image using FSL FIRST (Patenaude et al., 2011). Cerebral white matter ROIs, including left and right hemispheres only (thresholded at 80%), were also included.

### Data observations and postlabelling delay suggestions

Trends between mean grey matter CBF and ATT versus age and time-of-day (**Figure 2**A,C) were visualized using a sliding-window mean and standard deviation (window size 5 years/2.6 hours, respectively). A voxelwise map of the age association (**Figure 2**B) was generated by calculating the correlation coefficient between the CBF or ATT in each voxel in standard space (after quantile normalization) against subject age.

The ATT distribution (**Figure 3**A) was plotted as the cumulative histogram of all voxels in standard space within a grey matter mask (partial volume estimate of more than 70%) across all subjects. Postlabelling delay (PLD) suggestions (**Figure 3**B) were made by calculating the 95/99^th^ percentile ATT within the standard space grey matter mask for each subject, then determining the 95/99^th^ percentile of these values within a sliding window across ages (window width 5 years). This yields an estimate of the PLD required to accurately measure CBF with single-PLD ASL in 95/99% of voxels in 95/99% of participants.

### Deconfounding

In addition to a number of previously identified confounding factors for UK biobank brain imaging data (Alfaro-Almagro et al., 2021), including age, sex, time-of-day, site, head size, hardware/software changes, subject motion, table position and others, in this study we also incorporated the effect of haematocrit (known to affect blood T1 and therefore the measured ASL signal (Lu et al., 2004)) and cortical thickness, as a proxy for partial volume effects which are likely in many voxels and brain regions, especially given the relatively large voxel size used here. The mean haematocrit value across all available measurements and the mean cortical thickness across both hemispheres generated by the structural image analysis were split by site, as recommended previously (Alfaro-Almagro et al., 2021), and incorporated into the confound matrix. For the small number of subjects without haematocrit data (322), these values were set to the median of the other subjects.

All IDPs in this cohort of 7,157 subjects were processed for outlier removal (more than five times the median absolute deviation from the median) and quantile normalization prior to the confounds being regressed out of the data, as per previous studies (Miller et al., 2016; Wang et al., 2022). A comparable process was performed for voxel-wise CBF and ATT measures in standard space.

### Associations between ASL and other measures

As well as imaging data, thousands of variables are available in the UK Biobank dataset, including some derived from genetics data (e.g. polygenic risks scores and genetic principal components), lifestyle factors, physiological measurements (e.g. blood pressure), cognitive tests and health data. These factors were pre-processed using FSL’s FUNPACK (McCarthy, 2023) to convert each measure to a meaningful numeric value and then processed similarly to the IDPs for outlier removal, quantile normalization and deconfounding. Univariate associations between the 50 ASL IDPs and the 27,176 non-imaging factors were calculated using Pearson correlation. Some of these non-imaging variables have a significant amount of missing data, so we assessed the significance of the associations by the P values, which account for the degrees of freedom in the analysis. Multiple comparisons correction to give P_corrected_ < 0.05 resulted in a −log_10_(P_uncorrected_) threshold of 7.43 using the conservative Bonferroni method, or 5.11 using the False Discovery Rate (FDR) approach (Benjamini and Hochberg, 1995).

For those non-imaging measures that had at least one significant association (exceeding the FDR threshold) with an ASL IDP, the voxelwise correlation between this non-imaging measure and the deconfounded CBF and ATT maps was also calculated to allow examination of the spatial patterns associated with this relationship. This helps in the interpretation of the association, revealing more subtle features than are captured by the coarse ROIs used to generate the IDPs, including the identification of correlations that may be driven by confounding factors (e.g. a spatial pattern with strong correlations at the brain edges, which may be driven by partial volume effects or registration issues).

To more robustly investigate associations with selected binary health-related measures (depressive episode [ICD10 code F32.9] and self-reported hearing loss at the imaging visit [UK biobank data field 2247, instance 2]), the two groups of subjects were compared using permutation testing (Winkler et al., 2014) with threshold-free cluster enhancement (Smith and Nichols, 2009), to give greater confidence in interpreting the spatial patterns of these subtle correlations. Regions where the family-wise error corrected P value was less than 5% were considered significant.

### Assessing the sensitivity of ASL relative to other imaging measures

After finding significant correlations between ASL IDPs and some non-imaging measures, we wanted to establish whether other brain imaging modalities were equally as sensitive. Rather than just comparing the most significant ASL association with all other IDPs, which could suffer from a circularity issue, we instead compared association strengths found with groups of IDPs from different imaging modalities (which were processed as described below). For each non-imaging measure, the 90^th^ percentile −log_10_(P) value within each IDP group was calculated as a measure of the strength of the associations with that group as a whole. Permutations of the group labels were then performed to build a null distribution under the assumption that all IDP groups were equally sensitive to this non-imaging measure. The P value associated with the ASL group belonging to this null distribution can then be calculated.

Since the 90^th^ percentile −log_10_(P) value from each IDP group will have higher variability from smaller groups of IDPs, imaging modalities with more than 50 IDPs were split into subgroups of 50 or less and the ordering of these subgroups were also randomly chosen for each of 500 permutations, giving a conservative estimate of the P value associated with the ASL group.

### Correlations with other imaging measures

IDPs from other modalities were processed in a similar manner to the ASL IDPs, with outlier removal, quantile normalization and deconfounding (using the standard set of confound variables (Alfaro-Almagro et al., 2021)). Univariate correlations between these 3,633 IDPs (excluding cortical thickness, which had already been used as a confound variable for ASL) and the ASL IDPs were then calculated. Significance thresholds were calculated as per the non-imaging comparison, giving a −log_10_(P_uncorrected_) threshold of 6.56 or 3.21 using Bonferroni or FDR, respectively. Where a non-ASL IDP was found to be significantly associated with an ASL IDP, spatial maps of these correlations were also calculated.

## Data Availability

All imaging data (including raw images, derived maps and IDPs), phenotypes and genetics data are made available by UK Biobank via their standard data access procedure (see http://www.ukbiobank.ac.uk/register-apply).

## Data and Code Availability

A subset of the UK Biobank brain imaging data which included ASL in the protocol from the October 2023 release (7,157 participants) were used in this study. Permission to use this data was obtained via material transfer agreement as part of Data Access Application 8107. All imaging data (including raw images, derived maps and IDPs), phenotypes and genetics data are made available by UK Biobank via their standard data access procedure (see https://www.ukbiobank.ac.uk/use-our-data/apply-for-access/).

The mean CBF and ATT maps across all subjects in MNI space are available here: https://www.fmrib.ox.ac.uk/ukbiobank/index.html <TO be uploaded upon publication>.

Code to run the full UK Biobank image processing pipeline, including the ASL analysis, is freely available (https://git.fmrib.ox.ac.uk/falmagro/uk_biobank_pipeline_v_1.5). Code to generate the ASL-specific ROIs, process the ASL IDPs and voxelwise data and to generate the results presented in this paper are available here <LINK to be added upon publication>.

## Acknowledgements

This research was conducted using the UK Biobank Resource under application number 8107. We are very grateful to UK Biobank for making this dataset available and to all the study participants who kindly donated their time to make this resource possible. Thanks to Flora Kennedy-McConnell for help with generating the vascular territory regions of interest and to Niels Oesingmann and Federico von Samson-Himmelstjerna for help setting up the imaging protocol. TWO is supported by a Sir Henry Dale Fellowship jointly funded by the Wellcome Trust and the Royal Society (220204/Z/20/Z). SMS and KLM are supported by the Wellcome Trust (202788/Z/16/Z, 224573/Z/21/Z, 215573/Z/19/Z). MAC and MC were supported by the Engineering and Physical Sciences Research Council (EP/P012361/1 and EP/P012361/2), and the NIHR Biomedical Research Centre Nottingham. DLT was supported by the National Institute for Health and Care Research University College London Hospitals Biomedical Research Centre (NIHR UCLH BRC) and the Wolfson Foundation (PR/ylr/18575). The Wellcome Centre for Integrative Neuroimaging was supported by core funding from the Wellcome Trust (203139/Z/16/Z) with additional support from the NIHR Oxford Health Biomedical Research Centre (NIHR203316). Computation partially used the Oxford Biomedical Research Computing (BMRC) facility, a joint development between the Wellcome Centre for Human Genetics and the Big Data Institute supported by Health Data Research UK and the NIHR Oxford Biomedical Research Centre. The views expressed are those of the authors and not necessarily those of the NIHR or the Department of Health and Social Care. For the purpose of open access, the author has applied a CC BY public copyright license to any Author Accepted Manuscript version arising from this submission.

## Declarations

PMM gratefully acknowledges personal support from the Edmond J Safra Foundation. He has received consultancy fees from Mycobalan Therapeutics, Biogen, Astex, GlaxoSmithKlein, Sudo, Novartis and Nodthera over the course of this work. MAC and MC work for and hold equity in Quantified Imaging.

## Supplementary Figures

**Supplementary Figure 1:**
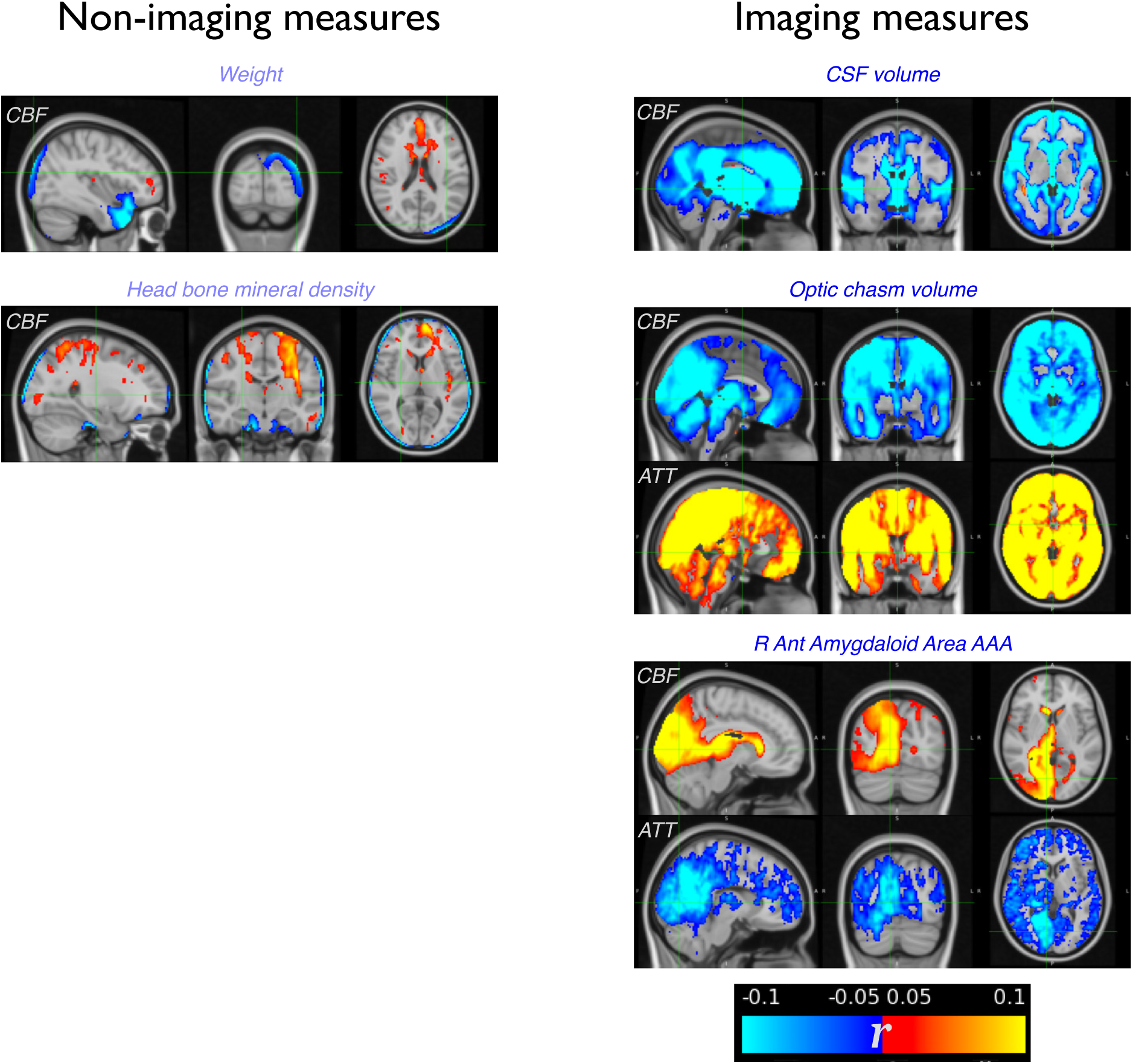
Examples of spatial maps where correlations with ASL IDPs may be artefactual. The smoothly varying CBF reductions at the brain edges that are associated with weight and bone mineral density could be driven by differences in magnetic field distortions or the ability of the registration algorithms to tackle variations in these physical factors. CSF volume likely drives partial volume effects, leading to apparent reductions in CBF adjacent to regions containing CSF. The optic chiasm is adjacent to brain feeding arteries, so this association is likely driven by artery morphology. Similarly, R Ant Amygdaloid Area AAA is a measure made adjacent to the right posterior cerebral artery, so estimation of this area is likely correlated with the artery diameter, leading to an apparent association in the vascular territory of this artery.

**Supplementary Figure 2:**
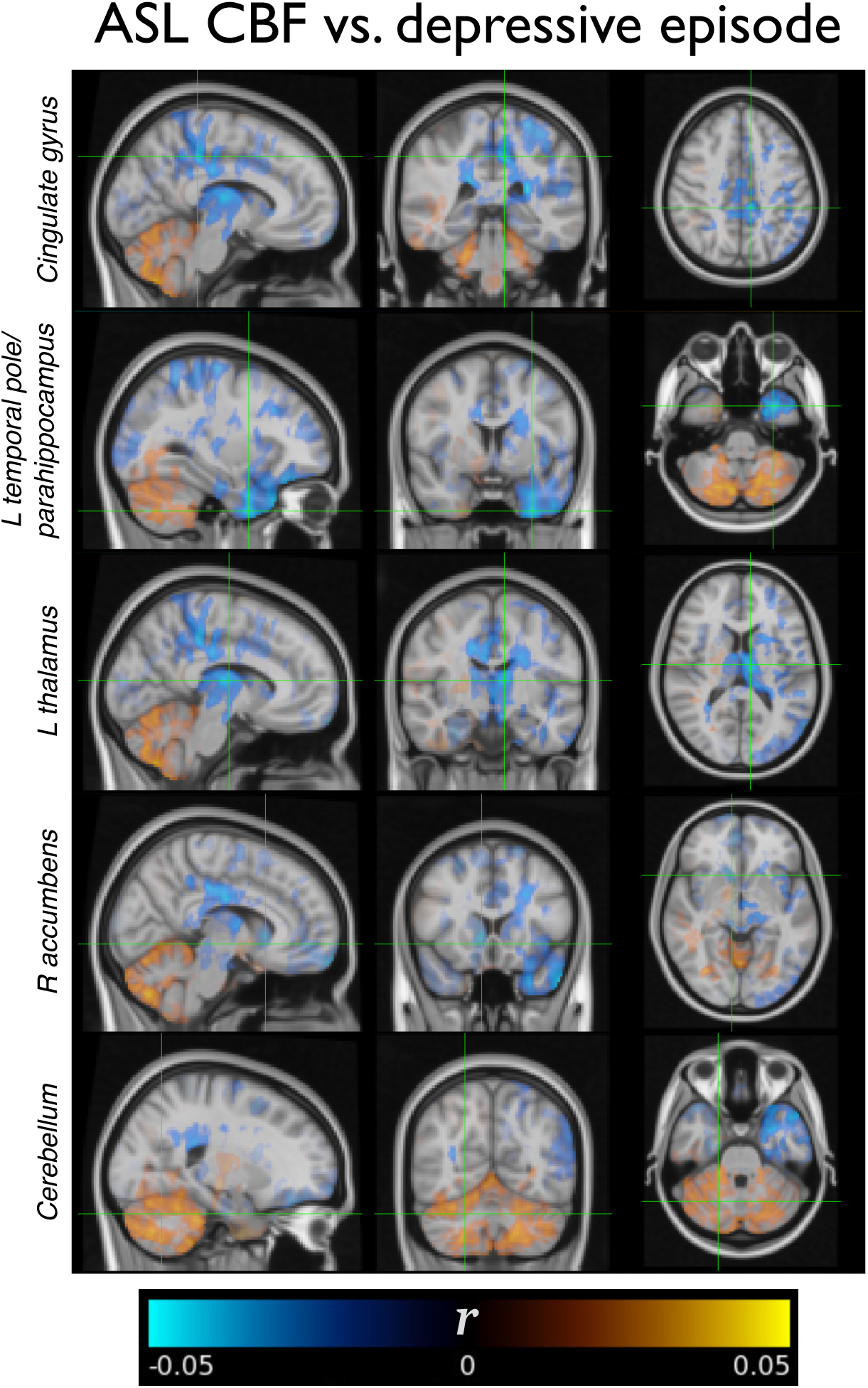
Additional spatial maps demonstrating the association between CBF and depressive episode. Whilst there are no significant clusters, regions showing higher correlations have previously been found to have altered connectivity in depression (Helm et al., 2018).

## Supplementary Table Captions

**Supplementary Table 1**: All significant associations between ASL IDPs and non-imaging measures passing the false discovery rate (FDR) threshold for multiple comparisons. Where more than one ASL IDP was associated with the non-imaging measure, only the most significant association is shown.

**Supplementary Table 2**: Non-imaging measures which were significantly more highly correlated with the ASL IDPs as a group, compared to other modalities.

**Supplementary Table 3**: All significant associations between ASL IDPs and other brain imaging IDPs passing the false discovery rate (FDR) threshold for multiple comparisons. Where more than one ASL IDP was associated with the other brain imaging IDP, only the most significant association is shown.

